# Transition from growth to decay of an epidemic due to lockdown

**DOI:** 10.1101/2020.12.14.20248154

**Authors:** H. Khataee, J. Kibble, I. Scheuring, A. Czirok, Z. Neufeld

## Abstract

We study the transition of an epidemic from growth phase to decay of the active infections in a population when lockdown measures are introduced to reduce the probability of disease transmission. While in the case of uniform lockdown a simple compartmental model would indicate instantaneous transition to decay of the epidemic, this is not the case when partially isolated active clusters remain with the potential to create a series of small outbreaks. We model this using a connected set of stochastic susceptible-infected-removed/recovered (SIR) models representing the locked-down majority population (where the reproduction number is less than one) weakly coupled to a large set of small clusters where the infection may propagate. We find that the presence of such active clusters can lead to slower than expected decay of the epidemic and significantly delayed onset of the decay phase. We study the relative contributions of these changes to the additional total infections caused by the active clusters within the locked-down population. We also demonstrate that limiting the size of the inevitable active clusters can be efficient in reducing their impact on the overall size of the epidemic outbreak.

**Statement of Significance:** Restricting movement and interaction of individuals has been widely used in trying to limit the spread of COVID-19, however, there is limited understanding of the efficiency of these measures as it is difficult to predict how and when they lead to the decay of an epidemic. In this article, we develop a mathematical framework to investigate the transition to the decay phase of the epidemic taking into account that after lockdown a large number of active groups remain with the potential to produce localised outbreaks affecting the overall decay of infections in the population. Better understanding of the mechanism of transition to the decay of the epidemic can contribute to improving the implementation of public health control strategies.

## 1 Introduction

Quantitative epidemiological models are being used as a tool to inform public health decisions about infectious disease outbreaks, like recently during the COVID-19 pandemic. Simpler mathematical models were useful to introduce concepts such as, the basic reproduction number *R*_0_ [1], while more complex agent based models were used to estimate hospital occupancy, or the effect of planned social distancing measures [2–4]. Arguably the simplest and most influential epidemic model operates with the susceptible, infected and removed/recovered subpopulations and hence referred to as the SIR model [5]. The corresponding system of nonlinear ordinary differential equations exhibits an initial exponential spread of an epidemic, followed by saturation as the susceptible proportion of the population decreases. Eventually, as the immune sub-population reaches a critical threshold, often referred to as herd-immunity threshold, the infected subpopulation decays to zero.

The simplest SIR model assumes that the epidemic parameters (e.g., the probability of transmission from an infected to a susceptible individual) remain constant during the outbreak, and the whole population is perfectly mixed: any two individuals have the same chance for disease transmission. However, in the COVID-19 pandemic a range of public health measures were introduced with the goal of reducing transmission efficiency. Accordingly, the initial exponential growth phase of the infectious fraction of the population was followed by an exponential decay after the introduction of social distancing measures, primarily reflecting the reduced transmission rate well before a significant fraction of immune population would have accumulated [6].

Detailed analysis of epidemic and mobility data from various European countries during the first wave of the COVID-19 pandemic has revealed that after the introduction of social distancing measures the transition from growth to decay phase may involve a fairly long, and country-specific delay [6]. The 4-5 days long incubation period of the disease [7, 8], as well as the average survival time of patients with fatal COVID-19 illness are expected to introduce a certain delay between the alteration of social behavior and its observable effect in epidemic data such as the daily new infections and death rate. However, in most countries the delay between the introduction of control measures to the peak of the epidemic was significantly longer, usually lasting for several weeks. The same study also revealed that the delay was longer in countries where the control measures were less strict [6].

Within the context of an SIR model a substantial sudden reduction of the infection transmission rate due to social distancing efforts should flip the exponential growth into exponential decay instantaneously. In this paper we demonstrate that the delay *τ* between the introduction of social distancing measures and the onset of the decaying phase of an epidemic can arise when control measures affect various groups of society highly selectively. For example, under official lockdown orders transmission can be reduced for the majority of the population, while the disease remains able to spread among various distinct clusters – like workers in warehouses, meat processing plants or among prison inmates. To study this effect we propose a generalization of the SIR model to take into account the heterogeneity of the population with respect to epidemic control measures. We consider a dual population structure where most of the population has a strongly reduced rate of transmission, while the transmission rate remains high within distinct clusters of a certain size. By stochastic simulations we explore how the delay *τ*, the decay rate and the total number of infected people are influenced by the size of the non-restricted sub-population as well as the typical size of the clusters within the non-restricted population.

## 2 The Model

To model the spread of the epidemic in a population we use the standard SIR epidemic model, defined by the following system of ODEs [5, 9]:

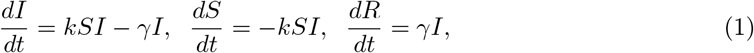

where *I, S*, and *R* denote the infected, susceptible, and recovered fractions of the population, respectively, i.e., *S* +*I* +*R* = 1. Here, *k* is the rate of transmission from *I* to *S* per infected individual, while *γ* represents the rate of recovery from *I* to *R*, either through acquired immunity or death. The behavior of the model is determined by a single non-dimensional parameter, the reproduction number *R*_0_ = *k/γ*, which gives the number of new infections caused by a single infected person within a fully susceptible population. The condition for an epidemic outbreak is *R*_0_ > 1, while for *R*_0_ <1 the epidemic dies out.

In the initial phase of an epidemic described by the SIR model most of the population is susceptible and the number of infected individuals grows exponentially. We are interested in the epidemic dynamics within a population under social distancing measures that have been implemented sufficiently early (i.e., *I, R* << 1 and *S* ≈ 1) and are efficient enough to reduce the reproduction number below one. In this regime, Equation (1) simplifies to *dI/dt* = *kI* − *γI* and yields:

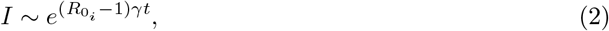

where parameters 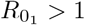 > 1 and 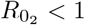 <1 characterise the initial growth and post-intervention decay phases of *I*, respectively.

To model the heterogeneity of the population with respect to epidemic control measures, we divide the total population of size *T* into sub-populations: (i) the locked-down population fraction of size 0 <*L* <1, where the social distancing measures are effective and the infection transmission rate reduces (*R*_0_ <1), and (ii) the rest of the population (e.g., nursing home inhabitants, prison inmates, essential service providers) are sorted into partially isolated clusters, and within the clusters the infection transmission rate remains unaffected (*R*_0_ > 1). For simplicity, we assume that each cluster has the same size, *M*, a model parameter that reflects both the organization of social institutions as well as potential efforts to control the epidemic. The number of such clusters is *N*, and thus the size of the non-socially distancing sub-population is *NM* = (1 − *L*)*T*.

The locked-down sub-population and each cluster has its own SIR dynamics, which are also coupled as depicted in Fig. 1(A). Specifically, we define the transmission rates as follows. Within the locked-down sub-population (*L*), we define *k*_*L*→*L*_ = *R*_02_*γ*. Thus, in this sub-population, in the absence of external interactions the number of infections would decay exponentially. In contrast, for each cluster *j* = 1,.., *N*, we assume that the infection transmission rate within the cluster remains unchanged, i.e., 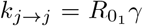. Transmission rates are also reduced between clusters and the locked-down sub-population: 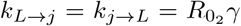. Finally, we also assume that there is no direct transmission of infection between different clusters.

**Figure 1:**
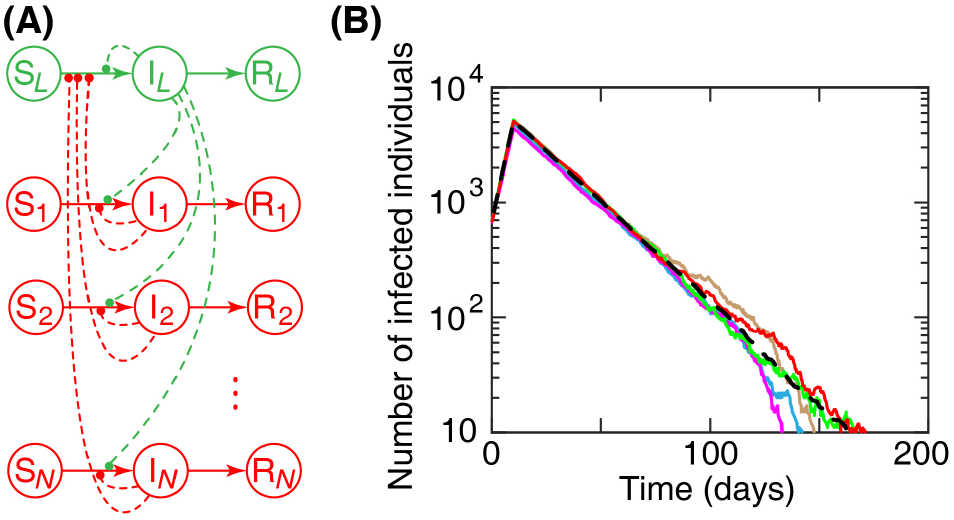
Susceptible-infected-removed/recovered (SIR) model. (A) SIR networks in the locked-down (green) and not socially distancing (red) sub-populations. The latter is composed of *N* clusters, each has its own SIR dynamics. *S, I*, and *R* are proportions of susceptible, infected, and recovered individuals, respectively. Dashed curves: infections transmitted by infected individuals to the susceptible individuals. (B) The total number of infected individuals versus time calculated using the Gillespie stochastic simulation algorithm. Solid lines: five example runs of the algorithm (in different colours) for *N* = 100 and *M* = 10. Black dashed line: *I*_tot_(*t*), average of 10 runs of the algorithm. Other simulation parameters are in Table 1.

We can now establish the dynamics for each of the SIR subsystems. For each cluster *j*, new infections occur at the rate given by:

**Table 1:** Model parameters.

| Parameter | Value |
| --- | --- |
| Total population size, $T$ | 1,000,000 |
| Initial reproductive number, $R_{0_1}$ | 3 |
| Reproductive number after lockdown, $R_{0_2}$ | 0.6 |
| Recovery rate, $\gamma$ | 0.1 ($\text{day}^{-1}$) |
| Initial ratio of infected individuals, $I_{0_1}$ | $667 \times 10^{-6}$ |
| Time of lockdown, $t_{\text{LD}}$ | 10 (days) |

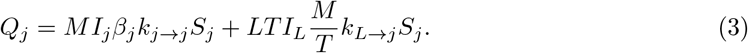

where *I*_*j*_ and *S*_*j*_ are the proportions of infected and susceptible in the cluster, the first term represents new infections transmitted within the cluster, and the second term describes infections introduced from the locked-down population. The parameter 0 <*β*_*j*_ <1 represents the probability that an individual in a cluster *j* interacts with others within her own cluster (we use *β*_*j*_ = 0.9). In the second term, the factor *M/T* captures the probability that an individual of the locked-down sub-population interacts with a member of cluster *j*.

Likewise, in the locked-down sub-population, the total rate of new infections is given by:

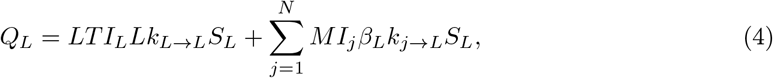

where *β*_*L*_ = 1− *β*_*j*_ = 0.1 represents the probability of interaction between individuals of cluster *j* and the locked-down sub-population.

Now, the dynamics of the proportion of infected individuals in the locked-down sub-population (*I*_*L*_) and in the active clusters (*I*_*j*_) are determined by the normalised rates of new infections combined with the rate of recovery, *Q*_*L*_*/LT* −*γI*_*L*_ and *Q*_*j*_*/M*− *γI*_*j*_, respectively. Note, while the SIR dynamics is the same for each cluster *j*, the temporal dynamics differs as the first in-cluster infection transmission could be initiated at various time points.

In the case of the standard SIR model it is natural to use a deterministic description, since in a large population the fluctuation of individual discrete events can be neglected and the overall dynamics is well approximated by continuous variables following mass action type kinetics. However, in our model, while the total population is still assumed to be large (for concreteness, we consider a city with total population *T* = 10^6^), after implementing lockdown strategies, the size of partially isolated clusters (where the infection is still able to propagate) are relatively small; we will assume typically values in the range *M* = 20 −150. Thus, a single new infected person in a cluster can increase *I*_*j*_ by a much larger extent than a similar infection in the larger population increases *I*_*L*_. Therefore, the discreteness of the population and the stochastic nature of the epidemic dynamics is important, especially at the initiation of epidemic outbreaks within the clusters, triggered by a single random infection event.

We implemented the time evolution of the interacting system of sub-populations as a series of discrete events in continuous time, by using the Gillespie stochastic simulation algorithm [10–12]. The parameter values used in the simulations are summarised in Table 1. We used 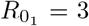 and 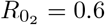 since the estimations of 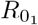 and 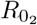 are in the ranges 2− 3.5 [13–15] and 0.36 − 0.82 [6], respectively. Although the exact value of *γ* for COVID-19 is uncertain [16, 17], we used *γ* = 0.1 (day*−*1) as a reasonable estimate for this parameter [18]. Yet, our analysis of the epidemic dynamics is rather general and does not rely on the values of the model parameters. Stochastic trajectories of the total number of infected individuals are shown in Fig. 1(B). The typical time course of the epidemic (*I*_tot_) was obtained as an average over 10 runs of the algorithm for each set of parameters *N* and *M*; see Fig. 1(B).

## 3 Results

We first explore how the time course of the total number of infected individuals in the whole population, *I*_tot_(*t*), is affected by the number and size of the clusters which remain active after the lock-down. We find that raising the number *N* and size *M* of these clusters can lead to a dramatic increase in the number of infections in comparison to the reference case when the whole population is locked down; dashed green line in Fig. 2(A-I). This difference is caused by a series of small outbreaks happening randomly when infection is transmitted into some of the active clusters while the overall level of infection decays in the rest of the population. The aggregate effect of the isolated outbreaks can be a delayed transition to the decaying phase of the epidemic relative to the time when the lockdown is implemented, in addition to a slower decay compared to what would be predicted based on the reproductive number 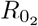 of the locked down population. Results in Fig. 2(A, D, G) also show that increasing the number of the active clusters has a negligible effect when the size of the clusters is small. These results suggest that dividing the active sub-population into groups with smaller sizes, independent of the number of groups, can reduce the total number of infected individuals in the population.

**Figure 2:**
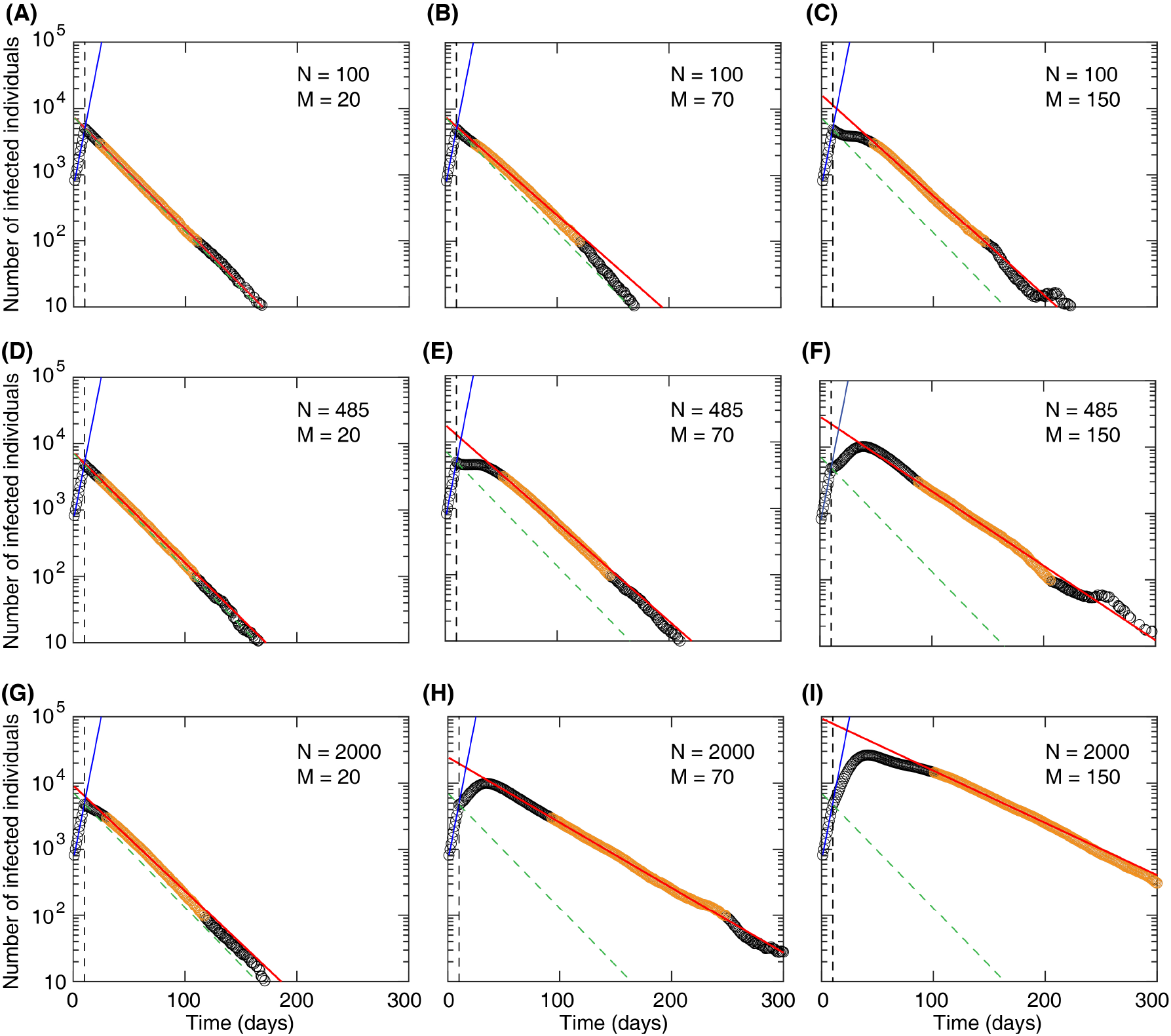
Total number of infected individuals (*I*_tot_(t)) versus time for different numbers (*N*) and sizes (*M*) of active clusters in the not socially distancing sub-population. Circles: *I*_tot_(*t*) calculated using the Gillespie algorithm after averaging over 10 repeated simulations. Solid red line is obtained by fitting 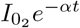 to the simulation data points highlighted by orange using the Mathematica routine NonlinearModelFit. To perform the fitting with minimal effects of the stochasticity, in most cases, the fitting range is set to start at a time after lockdown time *t*_LD_ where 100 <*I*_tot(*t*)_ <3000 (see Table S1 for the fit parameters 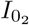 and *α*). Dashed green line: 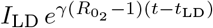 that passes through *I*_LD_ = *I*_tot_(*t*_LD_), where *t*_LD_ is the time of lockdown. Black dashed line: *t*_LD_. Blue line: 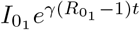. Other simulation parameters are in Table 1.

The significant differences observed in Fig. 2(A-I) in the number of infected individuals with varying the numbers and sizes of the active clusters in the population persuade us to further examine the transition to the decay of the epidemic. We characterise the transition by three properties: the decay rate of the number of infected individuals (*α*), the delay in the decay phase caused by the presence of the active clusters (*τ*), and the additional number of infected individuals (*I*_extra_) due to the slower and delayed decay of the epidemic.

The decay rate is calculated by fitting 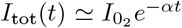 to the simulation data (averaged over 10 repeated realisations) within a selected range where there is a well defined exponential decay; see Fig. 2(A-I) and Fig. 3(A), orange circles. Then, the fit parameter *α* represents the decay rate (slope on the log-linear plot) of the number of infected individuals in the population. Least-squares fitting was performed using Mathematica (version 11, Wolfram Research, Inc.) routine Nonlinear-ModelFit.

**Figure 3:**
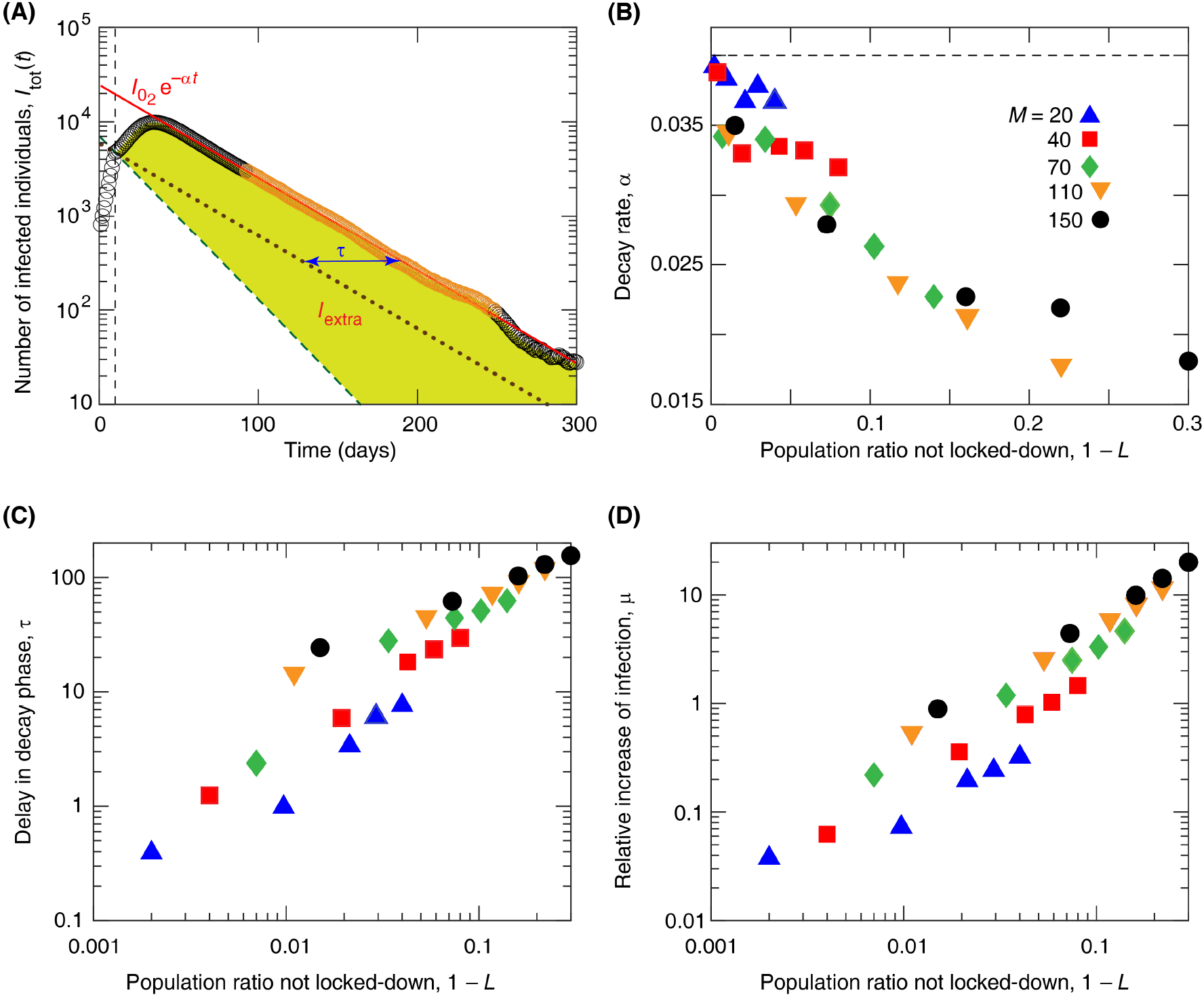
Analysis of the decay phase of an epidemic. (A) A sketch illustrating three proprieties of the decay phase (for example, with *N* = 2000 and *M* = 70). The decay rate *α* corresponds to the slope of the fit 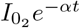 (red solid line). The delay *τ* in the epidemic decay, due to the presence of the active clusters, is the time-shift relative to an exponential decay with the same slope (*α*) that passes through the lockdown point (*t*_LD_, *I*_LD_) (brown dotted line); see Equation (5). Finally, the number of extra infected individuals *I*_extra_, calculated using Equation (6), is represented by the area of the yellow shaded region. (B-D) decay rate *α*, delay *τ*, and relative increase of total infections *µ* versus the active population ratio not locked-down, 1− *L*. Dashed line in (B): 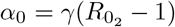. Other simulation parameters are in Table 1.

Next, the delay *τ* of the onset of the exponential decay phase, caused by the not locked-down clusters, is calculated as the time-shift relative to decay with the same rate *α* passing through *I*_LD_ = *I*_*tot*_(*t*_LD_) ≈ 5000; see Fig. 3(A). This decay follows 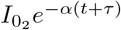 (illustrated using dotted brown line in Fig. 3(A)). Therefore, 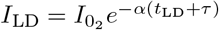 results in:

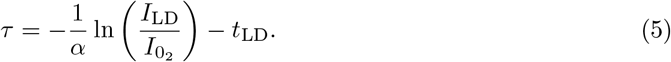

Finally, the number of new additional infected individuals, caused by the presence of not locked-down clusters, is calculated as:

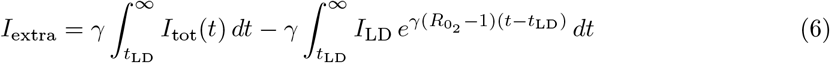

which corresponds to the area of the yellow shaded region in Fig. 3(A), multiplied by *γ*. Note that the second integral, representing the area under the dashed green line in Fig. 3(A), gives the number of individuals infected during the lockdown (*t* > *t*_LD_) if a complete lockdown was implemented with no active clusters (*N* = 0). This can be calculated explicitly and has the value 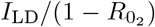, which we use as a reference value for the extra infections. Thus the effect of the clusters in the incomplete lockdown *N* > 0 can be characterised by the relative increase of total infections:

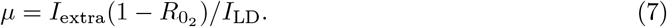

Results in Fig. 3(B) show that as the number and size of the active clusters increases the number of infected individuals decays slower than the value expected for a uniformly locked-down SIR model: *α*_0_ = *γ*(1− *R*_02_) = 0.04 (day^*−*1^). For the range of values considered in the simulations we find that when the locked-down fraction decreases to around *L* = 0.8 the characteristic decay time of the epidemic, *α*^*−*1^, increases from the reference value of 25 days (for *L* = 1) to around 50 days, i.e., *α* ≈ 0.02 (day^*−*1^).

Figure 3(C) shows that the delay of the decay phase, *τ*, can increase substantially in the range of tens to hundred days when the locked-down fraction is reduced to 80-90%. Also we find again that for a certain locked-down fraction, the delay can be much longer when the active population is composed of larger clusters. The relative increase of the total infections during the lockdown due to the active clusters (*µ*) shows a similar behavior (see Fig. 3(D)) indicating up to more than 10-fold increase when the active proportion is increased to around 10% of the population.

To further illustrate the differential impact of the number and size of the active clusters on the extra infections during the decay phase of the epidemic, we generate a heat-map representing *I*_extra_(*M, N*); see Fig. 4(A, B). Using a logarithmic scale for both variables *N* and *M*, the linear iso-contours in Fig. 4(B) suggest to approximate the dependence on these variables by a power law of the form:

**Figure 4:**
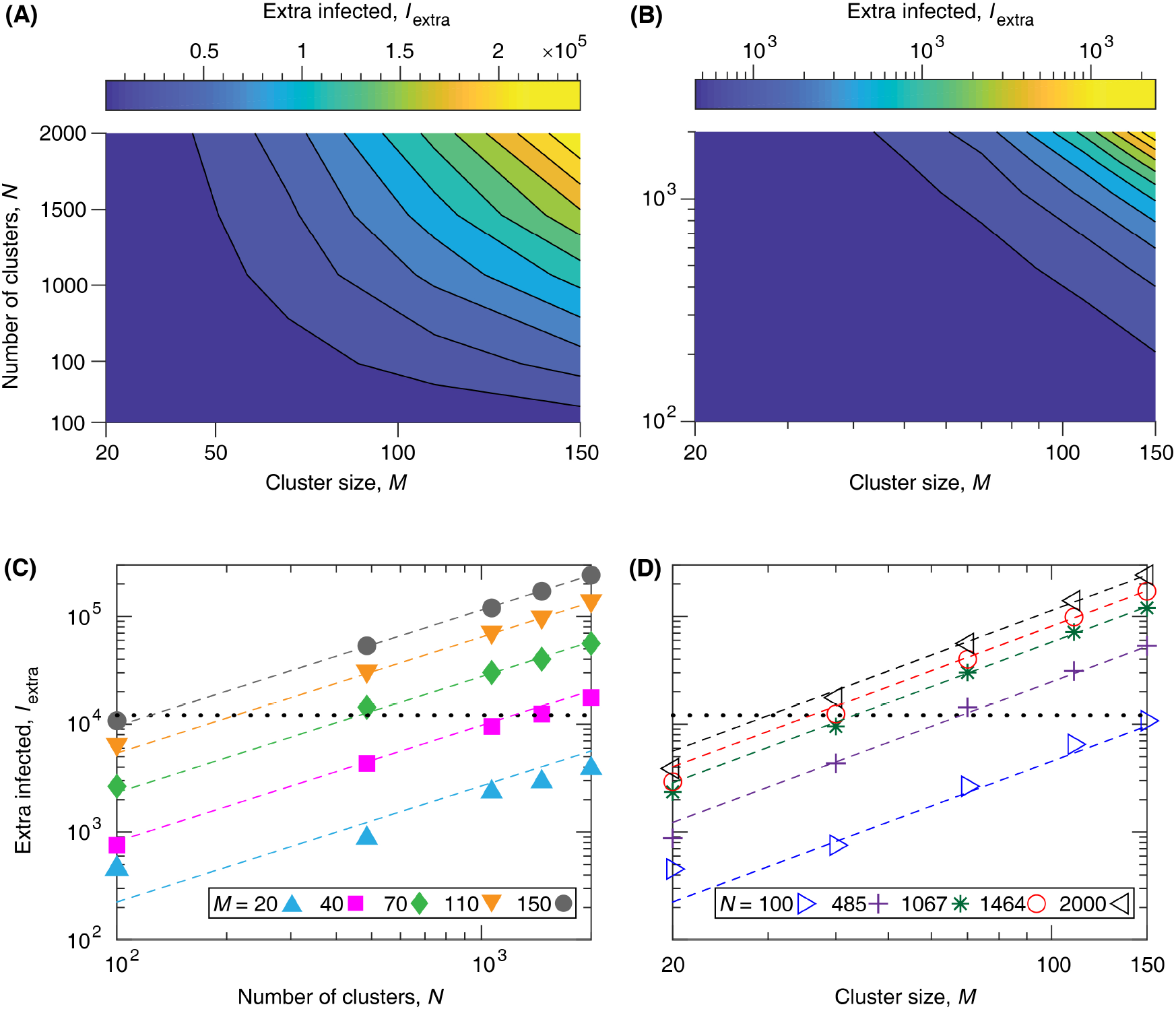
Analysis of the extra number of infected individuals, *I*_extra_ calculated using Equation (6). (A, B) *I*_extra_ versus *N* and *M*. (C, D) Dashed lines: fits *kN*^*a*^*M*^*b*^ to *I*_extra_(*N*; *M*) simulation data points, where *a* = 1.08 ±0.02, *b* = 1.86 ±0.02, and *k* = 0.01 ±0.001. Parameter values correspond to mean ± standard error (SE). Dotted lines: represent the reference value of total infected from the start of lockdown in a population without active clusters (*N* = 0), i.e., the area under the dashed green curve in Fig. 3(A).

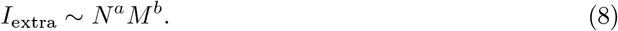

We find that this functional form leads to a good fit for the scaling of the data from the stochastic simulations (Fig. 4(C, D)) resulting in *a* ≈1 and *b*≈ 1.8. This confirms our earlier prediction that increasing the size of the clusters (*M*) has a stronger impact on the total infections than increasing the number of clusters (*N*) by the same proportion.

We further examine how the rate and delay of the decay phase contribute to the extra infections in the population. This relationship can be quantitatively expressed as:

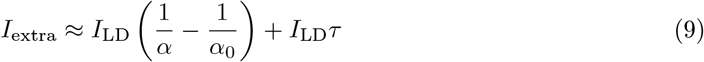

where the two terms represent the contributions due to slower and later decay, respectively. The approximation is based on replacing the transition region to a constant segment *I*(*t*) = *I*_LD_ until the onset of the decay phase (see S1(A)). Although this formula slightly underestimates the total infections in some cases, it provides a very simple representation of the two components.

We find a good agreement between the analytical and numerical calculations of the extra number of infected individuals; see Fig. S1(B), and the increase of infected individuals appears to be mainly due to the delay (*τ*) in the decay phase, rather than the rate (*α*) of the decay; compare Fig. S1(C, D). These results provide an explanation for our earlier prediction, that dividing the active sub-population into smaller clusters reduces the total number of infected individuals, mainly due to faster transition from growth phase to decay phase when the size of the outbreaks in infected clusters is smaller.

## 4 Discussion

We studied the effect of a large number of small active clusters within a locked-down majority population on the transition to the decay phase of an epidemic. We have shown that the active clusters can lead to delayed onset of the decay of the number of infected in the population and a slower decay rate of the epidemic. In our representative example simulations, considering a COVID-19-like epidemic in a city of one million inhabitants, we find that the delay can be in the range of several weeks to a few months while the characteristic time of the exponential decay, assumed to be around 3 weeks for the locked-down population, may double. This could lead to up to tenfold increase in the total number of infections occurring from the start of the lockdown relative to the case of a complete uniform lockdown. We found that the size of the clusters has a much stronger impact on the total extra infections in the incomplete lockdown than the number of these clusters, and the extra infections can be approximated with a double power law expression of the form: *I*_extra_ ∼ *N* · *M* ^1.8^. We also identified that the dominant factor leading to the extra infections is the delayed transition to the decay phase which has an approximately 5-fold larger contribution than the slower than expected decay.

The above results are based on a series of simplifying assumptions which allow us to keep a fairly simple general mathematical framework and efficient computer simulations while capturing the important impact of small minority groups in disease transmission, which is completely missed in standard ODE based compartmental epidemic models. This is particularly relevant in the context of the current COVID-19 pandemic, where unexpected strong over-representation of certain affected subgroups in the population become evident (e.g., very high proportion of nursing home deaths, or frequent outbreaks in meat factories, livestock plants, and prisons) [19–22]. Our model of course gives a very crude representation of these subgroups, considering a single average size of the clusters and assuming that the reproductive ratio remains the same as in the initial uncontrolled epidemic. Also, for simplicity, we assumed that the outbreaks occurring within the clusters are only stopped by the development of natural herd immunity within the cluster. In reality these outbreaks may be detected before they reach herd immunity and can be controlled through quarantine measures. This could be implemented quite easily in the model and would likely be equivalent to considering a smaller effective cluster size in the current model.

We also note, that the disproportionate impact of small sub-populations in epidemic models have also been observed in various other contexts. For example, in social contact network based epidemic models, it was shown that a very small fraction of highly connected individuals can have a large effect on the spread of infections in the population [23]. In the case of COVID-19, it was suggested that certain super-spreading events may play an essential role in the epidemic dynamics [24]. These examples also demonstrate potential limitations of mean field type models restricted to representing the average properties and behavior of the interacting components.

## Data Availability

This work is a mathematical model.

## Author Contributions

Z.N. and A.C. designed the research and developed the mathematical models. I.S. implemented the computational simulations. H.K. and J.K. analysed the modelling results. Z.N., H.K., A.C., and I.S. wrote the article.

## Supplemental Information

**Table S1:** Fit parameters 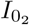 and *α* are approximated by fitting 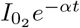 to the calculations of the average total number of infected individuals (highlighted by orange in Fig. 2). Parameter values correspond to mean ± standard error (SE).

| $N$ | $M$ | $I_{0_2}$ | $\alpha$ |
| --- | --- | --- | --- |
| 100 | 20 | 7515.69 ± 11.18 | 0.0392 ± 4 × 10<sup>-5</sup> |
| | 40 | 7647.35 ± 40.603 | 0.0388 ± 1 × 10<sup>-4</sup> |
| | 70 | 7709.89 ± 66.02 | 0.0342 ± 2 × 10<sup>-4</sup> |
| | 100 | 11372.40 ± 107.75 | 0.0344 ± 1 × 10<sup>-4</sup> |
| | 150 | 16005.40 ± 198.84 | 0.0350 ± 2 × 10<sup>-4</sup> |
| 485 | 20 | 7399.74 ± 20.67 | 0.0383 ± 1 × 10<sup>-4</sup> |
| | 40 | 8017.22 ± 96.69 | 0.0330 ± 3 × 10<sup>-4</sup> |
| | 70 | 18067.30 ± 187.33 | 0.0340 ± 2 × 10<sup>-4</sup> |
| | 110 | 24456.10 ± 167.05 | 0.0294 ± 1 × 10<sup>-4</sup> |
| | 150 | 36150.50 ± 476.94 | 0.0279 ± 1 × 10<sup>-4</sup> |
| 1067 | 20 | 8131.21 ± 40.50 | 0.0367 ± 1 × 10<sup>-4</sup> |
| | 40 | 12148.10 ± 81.04 | 0.0335 ± 1 × 10<sup>-4</sup> |
| | 70 | 23506.30 ± 125.86 | 0.0293 ± 1 × 10<sup>-4</sup> |
| | 110 | 34340.60 ± 329.14 | 0.0237 ± 1 × 10<sup>-4</sup> |
| | 150 | 61851.60 ± 953.33 | 0.0227 ± 1 × 10<sup>-4</sup> |
| 1464 | 20 | 9045.05 ± 69.96 | 0.0378 ± 2 × 10<sup>-4</sup> |
| | 40 | 14236.90 ± 111.06 | 0.0332 ± 1 × 10<sup>-4</sup> |
| | 70 | 23540.00 ± 119.12 | 0.0263 ± 1 × 10<sup>-4</sup> |
| | 110 | 42360.90 ± 541.74 | 0.0213 ± 1 × 10<sup>-4</sup> |
| | 150 | 103621.00 ± 1571.04 | 0.0219 ± 1 × 10<sup>-4</sup> |
| 2000 | 20 | 9252.42 ± 77.52 | 0.0367 ± 2 × 10<sup>-4</sup> |
| | 40 | 16975.60 ± 65.60 | 0.0320 ± 1 × 10<sup>-4</sup> |
| | 70 | 24625.70 ± 141.07 | 0.0227 ± 1 × 10<sup>-4</sup> |
| | 110 | 47621.70 ± 402.27 | 0.0178 ± 4 × 10<sup>-5</sup> |
| | 150 | 94487.40 ± 821.26 | 0.0181 ± 1 × 10<sup>-4</sup> |

**Figure S1:**
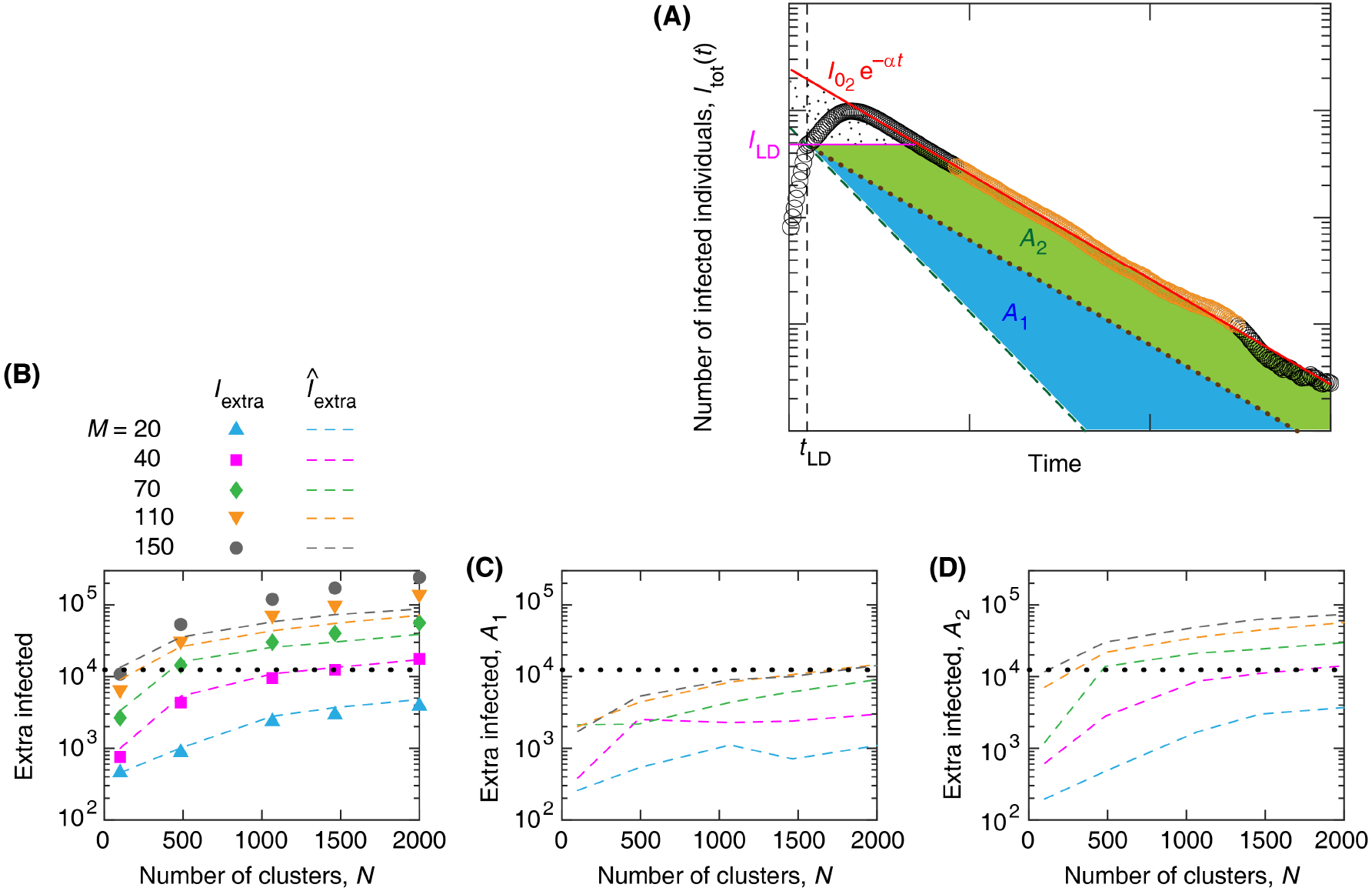
Contributions to the extra infected individuals, *I*_extra_ due to slower vs delayed decay of the epidemic. (A) A sketch illustrating areas associated with *I*_extra_, for example with *N* = 2000 and *M* = 70. Area *A*_1_ (blue shaded region) represents extra infected due to the slower rate of decay of the infections; first term in Equation (9). Area *A*_2_ (green shaded region) represents an estimate of the extra infected caused by the delay in the decay phase; second term in Equation (9). For clarity, the values for the *x* and *y* axes are not shown. (B) Comparison of the estimated extra infections (*Î*_extra_) calculated from Equation (9) and the numerically calculated *I*_extra_. Contributions to the extra infections due to slower decay (C) and delayed transition to the decay phase (D). Dotted line in (B-D) shows the total infections after the start of the lockdown if there were no active clusters (*N* = 0), i.e., the area under the dashed green curve in (A).

